# Exploring the Barriers and Facilitators of Vaccine Procurement in Health Districts of the South West Region of Cameroon

**DOI:** 10.64898/2026.11.09.26367572

**Authors:** Fai Bafon Theophile, Jerome Fru-Cho, Nkweleko Falone Fankam, Egbe Sarah Balle Tabi, Tah Aldof Yoah, Jane-Francis Tatah Kihla Akoachere

**Affiliations:** Department of Microbiology and Parasitology, University of Buea, Cameroon; Department of Public Health, University of Buea, Cameroon

**Keywords:** Barriers, Facilitators, Vaccine Procurement, Health Districts, South West, Cameroon

## Abstract

**Background:** Immunization is the most cost-effective way of preventing vaccine-preventable diseases and reducing morbidity and mortality in children 0 to 5 years old. The vaccine procurement capacity of health districts significantly influences the vaccine coverage rates. The advent of the socio-political crisis in the North West and South West Regions of Cameroon could have had a negative bearing on vaccine procurement and thus coverage rates. Thus, this study was carried out to explore the barriers and facilitators of vaccine procurement in health districts of a South West Region of Cameroon. This will help reduce vaccine stock-out at the health district level and increase vaccine coverage.

**Methodology:** A qualitative study was conducted using in-depth interviews (IDIs) to collect data from healthcare providers in all the 21 health districts in the South West Region working with the Expanded Program on Immunization (EPI), and who met the inclusion criteria. A Delphi consensus approach was employed to formulate feasible facilitators to the optimization of vaccine procurement. Data was analysed using Nvivo 11. Thematic analysis was used to explore the barriers to vaccine procurement at the level of health districts. A Delphi consensus of ≥ 70.0% was considered acceptable.

**Results:** From the thematic analysis, nine broad themes: financial constraints, limited workforce, inadequate transportation logistics, poor equipment, inadequate managerial skills, inadequate in-service training, vaccine stock-out and weak primary health care (PHC) system facilities emerged as barriers to vaccine procurement. Of the 12 strategies retained for the optimization of vaccine procurement, 5 (41.7%) were priority strategies: provision of extra solar refrigerators (100.0%), WHO certification training for EPI staff (100.0%), financial allocation for transport (100.0%), equipment and training, in-service training for both district and health facility staff (100.0%), resolving the socio-political crisis (100.0%) and provision of specialized transport logistics for maritime Health Districts (100.0%).

**Conclusion:** Financing of vaccine procurement, recruitment/certification of staff, provision of transportation logistics, provision of solar refrigerators, regular supervision, organization of in-service training, with focus on requisition/stock and bulk-requisition were the main strategies suggested to overcome the barriers to vaccine procurement.

**Recommendations:** These findings highlight the need for all stakeholders to co-finance and manage vaccine procurement at the health district level in the South West Region of Cameroon.

## INTRODUCTION

Immunization is recognized widely as one of the most successful and effective initiatives implemented for improving child survival globally [1]. The Expanded Program on Immunization (EPI), launched by the World Health Organization (WHO) in 1977 following the successful eradication of smallpox through mass immunization campaigns, has led to remarkable declines in the global incidence of many common vaccine preventable diseases including diphtheria, tetanus, pertussis, Hepatitis B, *Haemophilus influenzae* type B, and measles [1]. It is estimated that every year childhood vaccination prevents 2-3 million deaths worldwide and many more cases of illness and disability [2]. Despite these successes for childhood survival, there remain challenges to extending the benefits of immunization to all children. Childhood vaccination coverage in sub-Saharan Africa (SSA) is the lowest in World Health Organization regions. This region also experiences the highest burden of under-five deaths associated with infectious causes, many of which are vaccine preventable [3].

In Cameroon, the health system is yet to reach an optimal level of performance, as illustrated by the primary health indicators [4]. Indeed, despite a gradual decrease over the past decades, under-5 mortality rate in Cameroon remains unacceptably high. In 2020, under-five mortality was estimated at 72–82 per 1000 live births. In the same year, infant mortality rate (IMR) stood at 48–53 per 1000 live births [4]. The main causes of death in this paediatric population are malaria and conditions such as meningitis, acute lower respiratory tract infection, measles, diarrhea, and malnutrition [5]. To reduce childhood mortality, the government of Cameroon has developed and implemented several health development plans. For several decades, these plans focused on reducing childhood mortality via four interventions, notably: nutrition (breastfeeding, vitamin A supplementation), integrated management of childhood diseases, prevention of mother-to-child transmission of HIV and vaccination [6].

Vaccination is recognized as one of the most effective measures to prevent mortality, morbidity and complications from infectious diseases during childhood. It contributes to almost 25% of the reduction in infant and child mortality [7]. Over the past four decades, immunization has been at the center of Cameroon’s national public health strategies. Cameroon established its Expanded Program on Immunization in 1976 as a pilot, which went into full scale in 1982. At inception, the goal of the program was to reduce under five morbidity and mortality from vaccine preventable diseases (VPD) via the delivery of a full series of basic pediatric vaccines to eligible targets so as to provide the recipients with maximum protection against VPDs [8]. Since then, the Cameroon EPI has remarkably contributed to the reduction of infant morbidity and mortality, an achievement that clearly portrays a good program organization and delivery, relentless political commitment, equity and social justice [9].

Although far from achieving its ambitious goals, the EPI has made good progress in strengthening immunization service delivery and accelerating efforts to achieve VPD control over the past 40 years. This remarkable progress can partly be attributed to the support the program has been receiving from GAVI (Global Alliance for Vaccines and Immunization), The Vaccine Alliance, which has enabled the program to successfully introduce several new vaccines as well as scale up the use of old and underutilized vaccines [10]. Although Cameroon EPI registered a remarkable progress towards its immunization goals during the “Decade of Vaccines”, it is necessary to take stock of its achievements and advantage on its successes and lessons learned to inform a future strategy. This should be guided by existing evidence, epidemiology, policies and today social trends, which in turn will permit health authorities to consolidate the gains and face new challenges that may slow down the achievement of immunization targets of the Sustainable Development Goals (SDGs) [11]. To achieve this goal, the vaccine procurement capacity of health districts in Cameroon needs to be optimal.

The procurement chain for vaccines sourced from international donors involves a complex network of stakeholders, processes, and logistical challenges. From the initial order placement to the final delivery of vaccines to healthcare facilities, each step in the chain must be carefully monitored and evaluated to identify potential bottlenecks, risks, and areas for improvement. By establishing clear objectives and performance indicators for monitoring and evaluation, organizations can enhance transparency, accountability, and efficiency in the vaccine procurement process. By focusing on key areas such as timeliness, cost-effectiveness, quality assurance, transparency, compliance, risk management, stakeholder engagement, performance indicators, capacity building, and continuous improvement, organizations can optimize the procurement process and ensure that vaccines are delivered promptly. This study aimed at exploring the barriers and facilitators of vaccine procurement in health districts in the South West Region of Cameroon.

## MATERIALS AND METHODS

### Study design

A qualitative cross-sectional study design was employed to explore the barriers and facilitators of vaccine procurement among stakeholders of all the health districts of the South West Region of Cameroon. A Delphi consensus approach was used to develop an optimized model of vaccine procurement in the health districts.

### 3.2 Study area

This study was conducted in all the health districts of the South West Region of Cameroon (Figure 1). The South West Region (SWR) is one of the ten regions in Cameroon. The population of this region is estimated to be 1,762,752, with children under 5 years comprising about 42.9% [12]. By 2023, the region had 21 functional health districts, as documented by the Ministry of Public Health (MINSANTE). The map shows a representation of the health districts of the South West Region, newer districts (Toko and Bamusso) are yet to be updated into the map. The health district is the operational level of the health system. Each health district is headed by a District Medical Officer (DMO) who is closely assisted by a Chief Bureau Health (CBH). The coordinator of EPI heads vaccination at the District Health Service. All the 21 health districts are involved in vaccine procurement and have different vaccination coverage rates [12].

**Figure 1:**
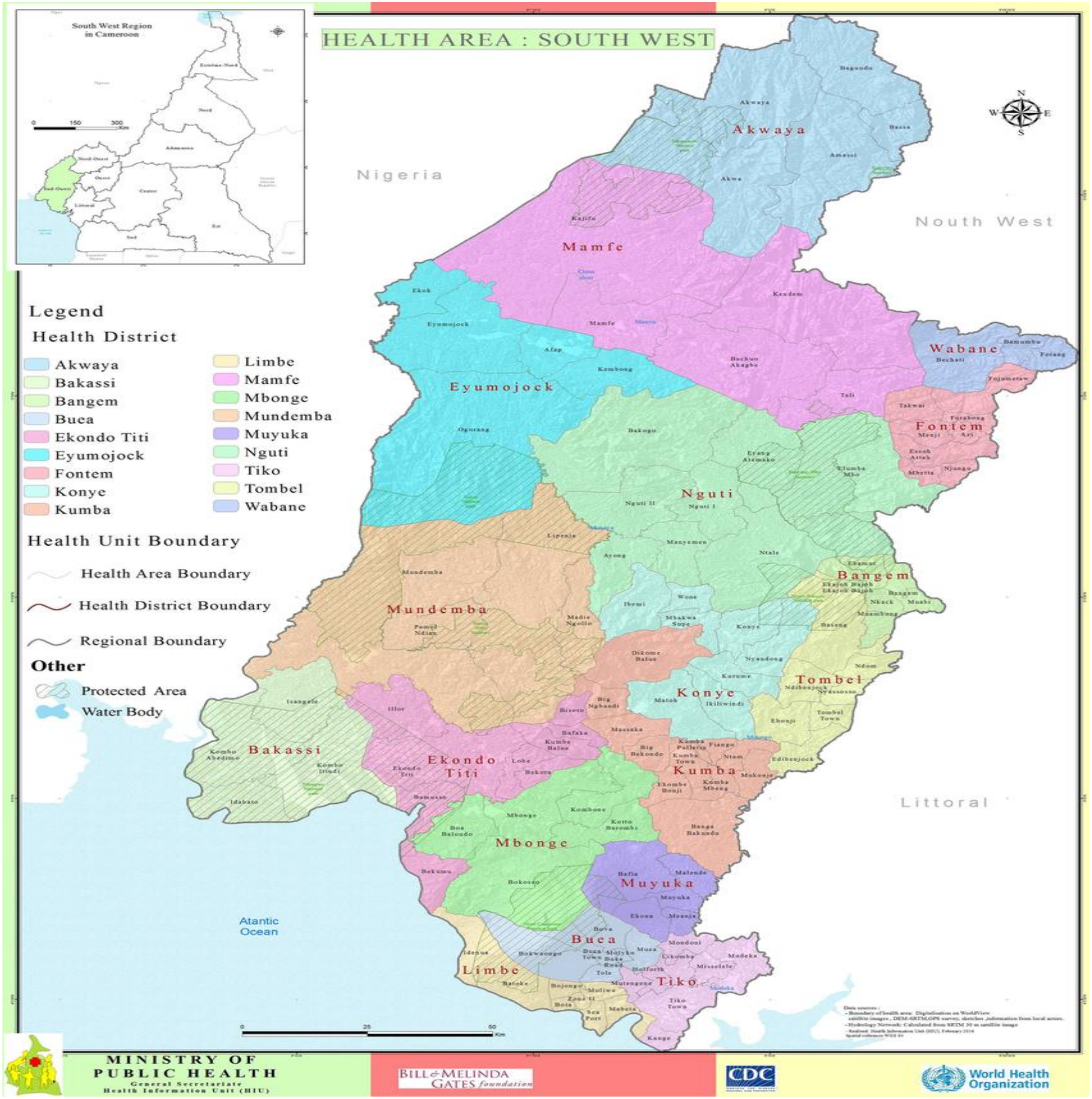
Health Map of the South West Region [13].

### 3.3 Study population

The study population or units for the in depth interview (IDI) and Delphi Consensus were regional and district healthcare providers involved in EPI for the SWR, Cameroon.

### Inclusion criteria

The study population or units consisted health care providers in charge of EPI at the district and regional level with at least two years of experience and granted consent to participate in this study.

### Exclusion criteria

Healthcare providers and other stakeholders who have not been involved in the vaccine procurement or EPI process for at least two years prior to this study,

### Sample size calculation

Healthcare providers and other stakeholders were interviewed until information saturation was reached.

### Sampling Technique

Regional and district EPI coordinators in SWR were purposefully sampled until information saturation point is reached.

### Data Collection Tool

An IDI guide was used to interview healthcare providers and other stakeholders with the aim of identifying the barriers and facilitators to vaccine procurement. The entire data collection tool consisted of two parts. Part one: IDI Guide –consisted of open-ended questions on the barriers and facilitators of vaccine procurement at the district level. Part two: Delphi Model development guide –consisted of a list of proposed solutions or measures aimed at mitigating the barriers to vaccine procurement.

### Training of the health professionals and field supervisors

Data collectors were health professionals including nurses and social workers. Training took three days and was aimed at equipping them with basic knowledge and skills for data collection and related procedures. The trainees were made familiar with the instruments for data collection. Training was conducted until the data collectors’ skill was found to be satisfactory. Only those who demonstrate adequate competence were engaged in the data collection process.

### Pre-Testing of Data Collection Tools

The data collection tools were pre-tested in Bamenda Health District in the North West Region of Cameroon and involved 05 healthcare providers and other stakeholders. This exercise helped to readjust some of the questions in order to increase validity of the data collection tools.

### Data Collection Procedure

IDIs were conducted with key EPI coordinators and other key stakeholders to explore the barriers and facilitators of vaccine procurement, and to develop a best-fit model for vaccine procurement. The interviews were recorded (with permission or consent from participants) and summary notes taken during the process. Data collection was carried out under close supervision of the principal investigator

### Data Management

For the qualitative study, the data was transcribed word verbatim and coded into themes. For Delphi model development, data was entered in an excel spreadsheet and coded.

### Data Analysis

To determine the barriers and facilitators of vaccine procurement, thematic analysis was used to identify the barriers and facilitators of vaccine procurement. Qualitative data was analyzed using Nvivo 11. IDIs were recorded, transcribed, and the texts were then reviewed to identify key themes deductively (informed by a review of the literature) and inductively (new themes emerging from the data collected). For the Delphi model development and IDIs, themes were used to develop the best model for the optimization of vaccine procurement at the level of the health district in the SWR of Cameroon. Themes were summarized in percentages and a Delphi consensus of ≥ 70.0% was considered acceptable.

### Ethical considerations

An ethical clearance was obtained from the Regional Ethics Committee for Human Research at the Regional Delegation of Public Health (Ref No480/CRERSH/SW/C/05/2025). Administrative authorisation was granted by the Regional Delegation of Public Health for South West, and District Health Services in the SWR. Consent to participate in the study was obtained from the healthcare providers and other stakeholders involved in the vaccine procurement chain.

## RESULTS

### Socio-demographic characteristics of key informants

The socio-demographic characteristics of 22 key informants purposefully selected to participate in this study are presented in table 1. The mean age of the key informants was 37.9 ± 1.43 years. Of the 22 key informants, the majority (9, 40.9%) were aged 30 – 39 years old and were males (10, 45.5%). Half (11, 50.0%) of them had at least a Masters in Public Health degree. The key informants were mainly Chief of Bureau Health (CBH) (59.1%). This was followed by Cold Chain Managers (18.2%), while Data Managers were the least (9.1%). Majority 10 (45.5%) of the key informants had work with EPI for 6 – 10 years.

**Table 1:** Socio-demographic Characteristics of key informants.

| **Variables** | **Frequency** | **Percent (%)** |
|---|---|---|
| **Age (in years)** |  |  |
| 20 – 29 | 03 | 13.6 |
| 30 – 39 | 09 | 40.9 |
| 40 – 49 | 07 | 31.8 |
| 50+ | 03 | 13.6 |
| **Gender** |  |  |
| Male | 10 | 45.5 |
| Female | 12 | 54.5 |
| **Professional Qualification** |  |  |
| Higher National Diploma /SRN | 02 | 9.1 |
| Bachelor of Science | 06 | 27.3 |
| Medical Doctor | 03 | 13.6 |
| Master in Public Health and above | 11 | 50.0 |
| **Duty Post** |  |  |
| Chief of Bureau Health | 13 | 59.1 |
| District Medical Officer | 03 | 13.6 |
| Data Managers | 02 | 9.1 |
| Cold Chain Manager | 04 | 18.2 |
| **Working Experience with EPI** |  |  |
| 1 – 5 years | 07 | 31.8 |
| 6 – 10 years | 10 | 45.5 |
| ➢ 10 years | 05 | 22.7 |

SRN-Sate Registered Nurses, EPI- Expanded Program on Immunization

### Barriers to vaccine procurement in health districts in the South West Region

Eight broad themes: financial constraints, limited workforce, inadequate transportation logistics, poor equipment, inadequate managerial skills, inadequate in-service training, vaccine stock-out and weak primary healthcare facilities emerged from the thematic analysis of the IDIs. These have been presented on Table 2 and further explained.

**Table 2:**
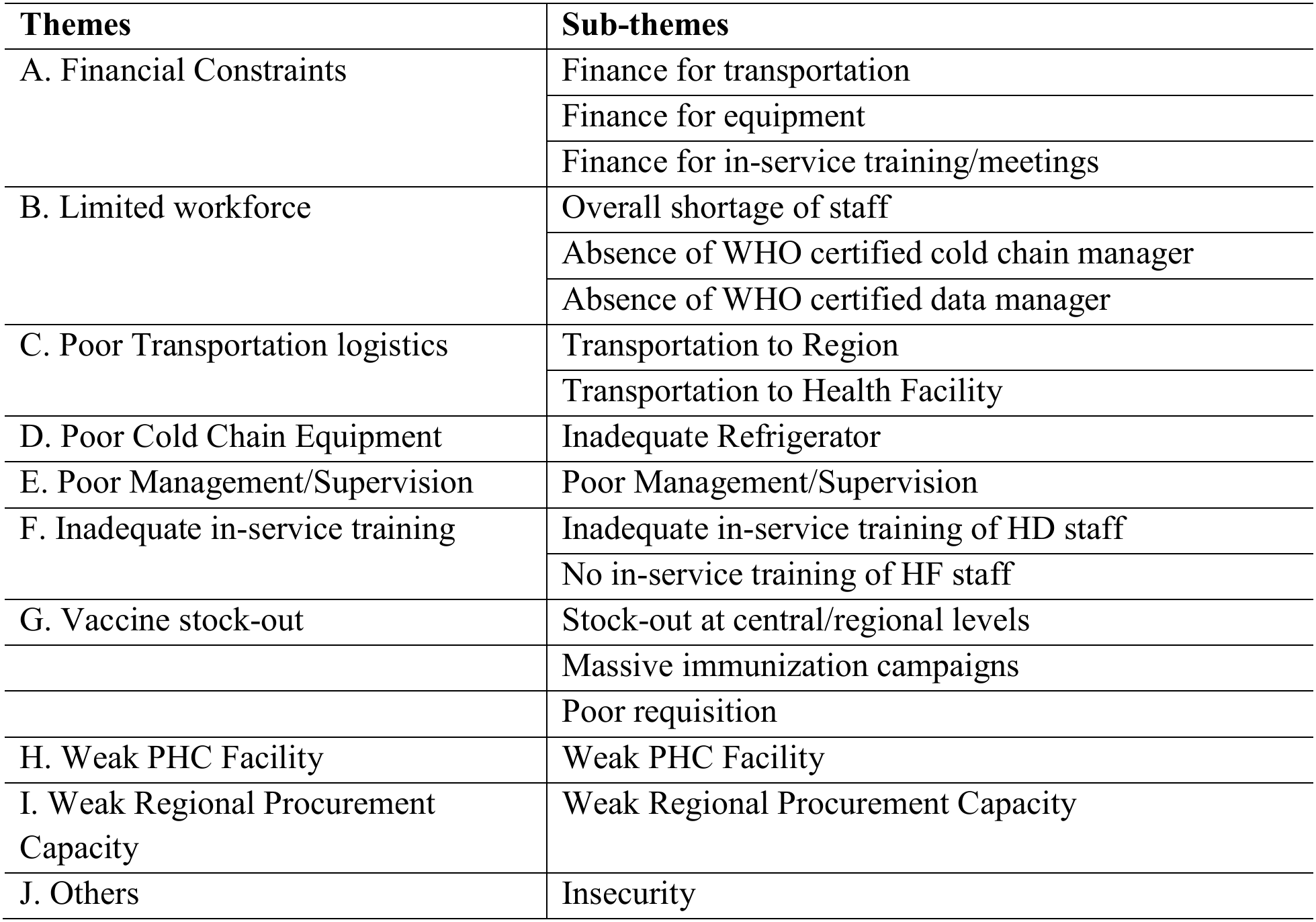
Barriers to vaccine procurement in health districts in the South West Region.

#### A. Financial Barriers

Majority of the participants said they encountered financial challenges in the course of vaccine procurement since this activity did not have direct funding. They indicated that there are usually no finances allocated to transport the vaccines from the region to the district. These views are captured in the quotes below:

> *“………because of the distance and topography of our health district, we usually have limited or no financing for the transportation of vaccines to the district and within it”*
>
> [Male Participant, in their 40s, Wabane HD, IDI].

Some participants in the IDI indicated that some leading health facilities lack finances to transport the vaccines from the district to the health facility. This could limit the timely distribution of the vaccines to remote areas. This opinion is presented below:

> *“ sometimes the leading health facilities complain that they lack money to come collect vaccines at the district. This causes stock-out and distorts the vaccine procurement process”*
>
> [Female Participant, in their 40s, Kumba North HD, IDI].

#### B. Limited Workforce

The majority of participants from the IDI (all age groups and genders) highlighted that staff shortage was a major hindrance to procurement of vaccines in the South West Region of Cameroon. Some health districts indicated lack of WHO certified cold chain manager and data manager. The absence of these professionals could lead to regular stock-outs and poor vaccine quality assurance. These assertions are captured below:

> *“………we have a chronic shortage of staff at the district level and this extends to EPI. The district does not currently have a cold chain manager talk less of a data manager. How do we expect vaccine procurement to be optimal”*
>
> [Male Participant, in their 50s, Tombel HD, IDI].

Some of the key informants went ahead to indicate that even the staff available lack the required competence in cold chain management and data validation for vaccines requisition. This opinion is explained below:

> *“ even the few staff that we have for EPI activities at the district do not have the required skills in cold chain and data management. This can be due to the fact that the district does not organize in-service training to improve staff skills in vaccine procurement activities”*
>
> [Male Participant, in their 30s, Ekondo Titi HD, IDI].

#### C. Inadequate Transportation Logistics

Some of the key informants observed that transportation logistics for vaccines from the region to the district and to the health facilities was grossly inadequate. This is because some districts have to come to the Region to collect their vaccines during the rainy season. Equally, maritime and unsafe districts have to come to safe areas to collect the vaccines. These observations are captured in the quotes below:

> *“ we are compelled to go Limbe or Buea to collect our vaccines because not everybody can take the engine boats to reach our districts. Given that, the maritime transport system here in Bakassi and Ndian is inadequate. We do understand this but finances should be allocated for this activity”* [Male Participant, in their 40s, Bakassi Titi HD, IDI].
>
> *“ during the rainy season, the Region expects us to come and collect the vaccines from Buea because the of the topography and bad roads that exist in Lebialem”*
>
> [Female Participant, in their 30s, Fontem HD, IDI].

Some of the participants from the IDI said there is no transportation logistic to facilitate leading health facilities to collect their vaccines at the level of the health district. This idea is captured in the statement below;

> *“ Most of the leading health facilities – Chief of Centers (COCs)are complaining that they have to come to the district to collect vaccines without any transportation logistics and financing put in place”*
>
> [Female Participant, in their 40s, Muyuka HD, IDI].

### D. Poor Cold Chain Equipment

A few key informants indicated that their cold chain was inadequate as they had faulty refrigerators. This could greatly affect the quality of vaccines sent out to the health districts. This opinion is highlighted below;

> *“………We had a refrigerator, but it has become faulty due to continuous voltage and power disruption. The present state of our refrigerator is not the best”*
>
> [Female Participant, in their 30s, Konye HD, IDI].
>
> *“ We have a solar powered refrigerator that works well during the dry season but occasionally does not function properly during the rainy season or when the sun fails to shine for several days”*
>
> [Female Participant, in their 40s, Kumba North HD, IDI].

#### E. Limited In-service training

Some of the key informants indicated that the in-service training on vaccine procurement was inadequate. There were only few training sessions per year (only one session on vaccine procurement in 2025). This could be due to the reduction in foreign aid. These views are captured in the quotes below;

> *“ we hardly have in-service training on vaccine procurement and EPI at the level of the Region and even when such information reaches us, we have work and financial constrains to go to the Region.”*
>
> [Male Participant, in their 50s, Bamusso HD, IDI].
>
> *“ a few in-service trainings are usually organized at the level of the Region but transportation consideration makes it difficult for us to leave Bakassi to Buea and back”*
>
> [Female Participant, in their 30s, Bakassi HD, IDI].

Majority of the key informants indicated that the few or no in-service training on vaccine procurement are organized in the health districts for chief of centers and health facilities. This implies that the relay of information or skills from the district level to the health facility level is inadequate. This observation is related in the statement below;

> *“ the district does not have the logistics and finance to organize in-service training for COCs and other health facility personnel on vaccine procurement”*
>
> [Female Participant, in their 40s, Buea HD, IDI].

#### F. Weak Primary Health Care

Some of the key informants indicated that leading health facilities do not come to collect their vaccines on time at the level of the health district. This could slow down vaccine procurement and cause unnecessary stock-outs and drop in vaccine coverage. This view is captured in the narrative below;

> *“………some COCs in our districts do not come for their vaccines on time. This slows down vaccine distribution and may result to poor vaccine coverage in the long run.”*
>
> [Female Participant, in their 30s, Tiko HD, IDI].
>
> *“ some COCs in our districts do not come for their vaccines on time; this causes unnecessary stock-outs and consequently slows down vaccine distribution and may result to poor vaccine coverage in the long run.”*
>
> [Female Participant, in their 20s, Mbonge HD, IDI].

#### G. Stock – Out of Vaccine

Majority of the key informants reported vaccine stock out especially for polio and HPV due to stock-out at the regional and central levels. This opinion is illustrated in the quote below;

> *“ the main reasons for stock-out in some antigens at the level of the health district is due to an upfront shortage at the regional and central levels. For example, we had a chronic shortage of HPV and polio because the region did have these antigens”*
>
> [Female Participant, in their 20s, Limbe HD, IDI].

Some of the key informants reported vaccine stock out especially for polio and HPV due to massive immunization campaigns that caused them to use up all the antigens. This observation is illustrated in the quote below;

> *“ one of the main reasons for stock-out in some antigens at the level of the health district is due to massive immunization campaigns beyond what the district had planned. For example, we had a chronic shortage of HPV because of such campaigns”*
>
> [Male Participant, in their 30s, Toko HD, IDI].

Some of the key informants reported vaccine stock out especially for polio and Penta due to poor requisition skills by EPI staff. This observation is illustrated in the quote below;

> *“ one of the reasons for stock-out in some antigens at the level of the health district is due to poor vaccine requisition skills by staff. For example, we had a chronic shortage of Rota vaccine because of such errors in requisition”*
>
> [Male Participant, in their 40s, Akwaya HD, IDI].

#### I. Poor Management/Supervision

Key informants reported that supervision from the regional level was grossly absent. This could be blamed on the security stakes of the region. The consequence is that the districts are left lose to do whatever pleases them. This opinion is illustrated in the quote below;

> *“………for the past 5-6 years I have not seen any team from the regional level coming to supervise any activity in the district. This can have a negative bearing on the functionality and operationality of district like ours”*
>
> [Male Participant, in their 40s, Wabane HD, IDI].
>
> *“ we have not had supervision from Buea (Regional Delegation) for a very long time. This can have a negative effect on vaccine procurement process, leading to stock out and low vaccine coverage”*
>
> [Female Participant, in their 50s, Toko HD, IDI].

#### H. Others – Insecurity

A few key informants indicated that security challenges prevented them from access to the region to collect vaccine and attend in-service training. This had a direct repercussion on vaccine stocks and coverage at the level of the health district. This narrative is captured in the sentence below;

> *“ since the beginning of the socio-political crisis, we take security precautions before going out to the region. Consequently, we have few antigens that are out-of-stock and our vaccine coverage has dropped significantly”*
>
> [Male Participant, in their 30s, Fontem HD, IDI].
>
> *“ since the beginning of the socio-political crisis, we only go out when it is safe. Consequently, we have vaccine shortages and quality assurance issues.”*
>
> [Male Participant, in their 30s, Eyomojock HD, IDI].

#### I. Inadequate Regional Vaccine Procurement Capacity

Majority of the key informants reported vaccine stock out especially for polio and HPV due to stock-out at the regional and central levels. This opinion is illustrated in the quote below;

> *“ the main reason for stock-out in some antigens at the level of the health district is due to an upfront shortage at the regional and central levels. For example, we had a chronic shortage of HPV and polio because the region did have these antigens”*

### Strategies to overcome the barriers to vaccine procurement in health districts in the South West Region

Eight broad themes; financing vaccine procurement, recruitment/certification of staff, provision of transportation logistics, provision of solar refrigerators, regular supervision, organize in-service training, training in requisition/stock and bulk-requisition, emerged from the thematic analysis of the IDIs. These have been presented on Table 3 and have been explained below.

**Table 3:**
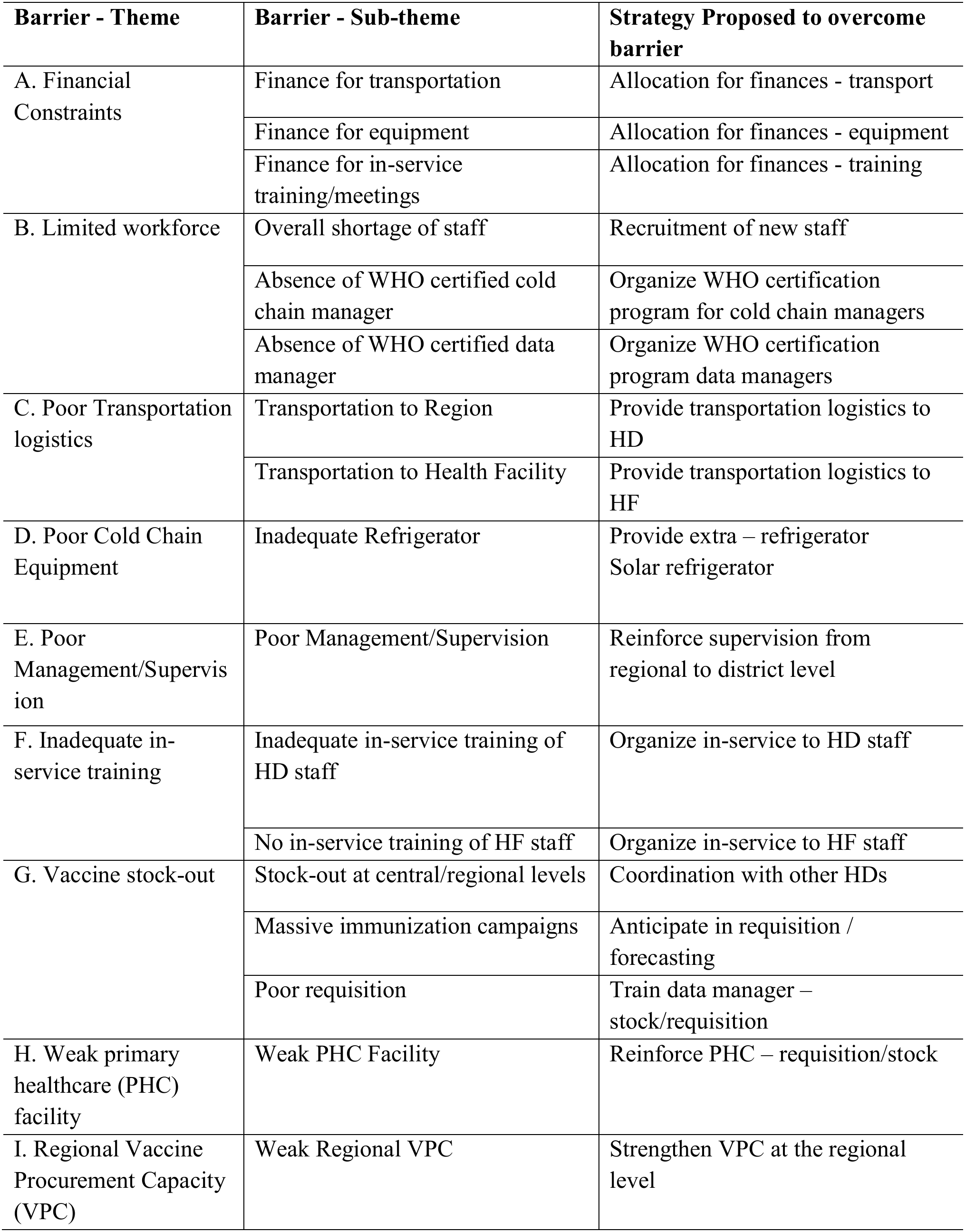
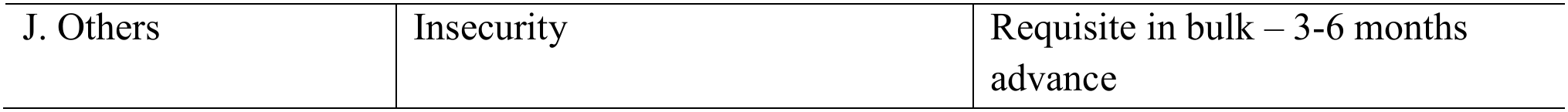
Strategies to overcome the barriers to vaccine procurement in health districts.

| **Barrier - Theme** | **Barrier - Sub-theme** | **Strategy Proposed to overcome barrier** |
|---|---|---|
| A. Financial Constraints | Finance for transportation | Allocation for finances - transport |
|  | Finance for equipment | Allocation for finances - equipment |
|  | Finance for in-service training/meetings | Allocation for finances - training |
| B. Limited workforce | Overall shortage of staff | Recruitment of new staff |
|  | Absence of WHO certified cold chain manager | Organize WHO certification program for cold chain managers |
|  | Absence of WHO certified data manager | Organize WHO certification program data managers |
| C. Poor Transportation logistics | Transportation to Region | Provide transportation logistics to HD |
|  | Transportation to Health Facility | Provide transportation logistics to HF |
| D. Poor Cold Chain Equipment | Inadequate Refrigerator | Provide extra – refrigerator<br>Solar refrigerator |
| E. Poor Management/Supervision | Poor Management/Supervision | Reinforce supervision from regional to district level |
| F. Inadequate in-service training | Inadequate in-service training of HD staff | Organize in-service to HD staff |
|  | No in-service training of HF staff | Organize in-service to HF staff |
| G. Vaccine stock-out | Stock-out at central/regional levels | Coordination with other HDs |
|  | Massive immunization campaigns | Anticipate in requisition / forecasting |
|  | Poor requisition | Train data manager – stock/requisition |
| H. Weak primary healthcare (PHC) facility | Weak PHC Facility | Reinforce PHC – requisition/stock |
| I. Regional Vaccine Procurement Capacity (VPC) | Weak Regional VPC | Strengthen VPC at the regional level |
| J. Others | Insecurity | Requisite in bulk – 3-6 months advance |

### Framework for the optimization of vaccine procurement in health districts in the SWR Results of Round One

Round 1 of the Delphi Model: Of the 25 participants who were invited to take part in the study, 22 participated in Round 1 (three of the invited participants did not commence Round 1) giving an 88.0% response rate). Participants’ ranking of outcome measures for Round 1 is provided in Table 4. Based on Round 1 ranking, 10 of the 12 outcomes for ‘strategies to optimize vaccine procurement’ combined met the consensus threshold of ≥70% respondents scoring an outcome from seven to nine and <15% scoring it one to three (combined consensus rate =83.3%). Those statements for which all respondents scored 7–9 and statements for which most respondents scored 1–3 are illustrated in Tables 5.

**Table 4:** Likert Scale Scores for ‘Strategies to optimize vaccine procurement’ Statements in Round 1.

| **Recommendation** | **Round 1** |  |  |  |
|---|---|---|---|---|
|  | **n** | **Score<br>1-3<br>n (%)** | **Score<br>4-6<br>n (%)** | **Score<br>7-9<br>n (%)** |
| Reinforce community involvement in vaccine procurement | 22 | 02(9.1) | 08(36.4) | 12(54.5) |
| Provision of extra solar refrigerators | 22 | 00(0.0) | 00(0.0) | 22(100.0) |
| Intensified training of staff in data and stock management | 22 | 02(9.1) | 05(22.7) | 15 (68.2) |
| Coordination between districts to reduce vaccine stock-outs | 22 | 02(9.1) | 02(9.1) | 18(81.8) |
| Recruitment of more EPI staff | 22 | 00(0.0) | 02(9.1) | 20(90.9) |
| WHO certification training for EPI staff | 22 | 00(0.0) | 00(0.0) | 22(100.0) |
| Bulk (3 months) requisition for hard to access health districts | 22 | 00(0.0) | 02(9.1) | 20(90.9) |
| Allocation of finance for transport, equipment and training | 22 | 00(0.0) | 00(0.0) | 22(100.0) |
| Regular supervision/monitoring from the region/central level | 22 | 00(0.0) | 02(9.1) | 20(90.9) |
| Provision of transport logistics to district to carry vaccines | 22 | 00(0.0) | 01(4.5) | 21(95.5) |
| In-service training for both district and health facility staff | 22 | 00(0.0) | 00(0.0) | 22(100.0) |
| Support health facilities in vaccine procurement | 22 | 01(4.5) | 03(13.6) | 18(81.8) |

### Results of Round Two

Twenty of the 22 participants of Round 1 (90.9%) took part in and completed Round 2 (Table 5). Round 1 participants suggested that five outcomes be added in Round 2, thus resulting in a new tally of 17 items for the combined measures. The 5 outcomes that were added to the ‘Strategies to optimize vaccine procurement’ were: resolving the sociopolitical crisis which is a key barrier to vaccine procurement, making provision for internally displaced persons (IDPs) during vaccine requisition, organizing regular evaluation meetings to monitor vaccine coverage, providing a specialized transport logistics for maritime HDs and strengthen vaccine procurement capacity at the region. Thirteen of the 17 outcomes (consensus rate= 76.5%) met the ≥ 70 and <15% criteria. For the 12 items that were also on Round 1, 08 outcomes (consensus rate= 66.6%) met the threshold criteria. Two items that met the criteria in Round 1 did not do so in Round 2. These items were ‘Support health facilities in vaccine procurement’ and ‘Coordination between districts to reduce vaccine stock-outs’. Meanwhile, all the 5 items that were added to Round 2, met the criteria.

**Table 5:**
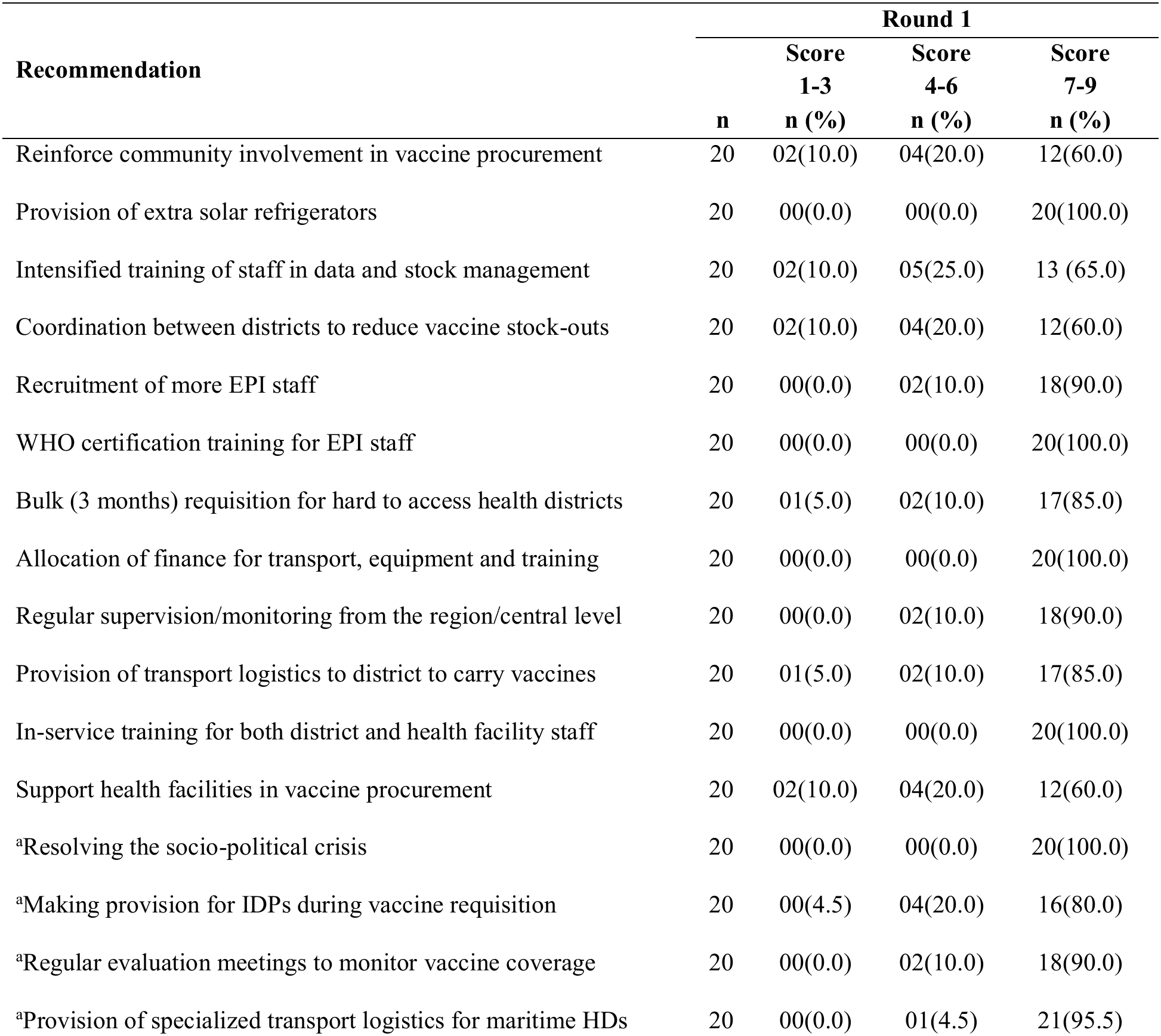
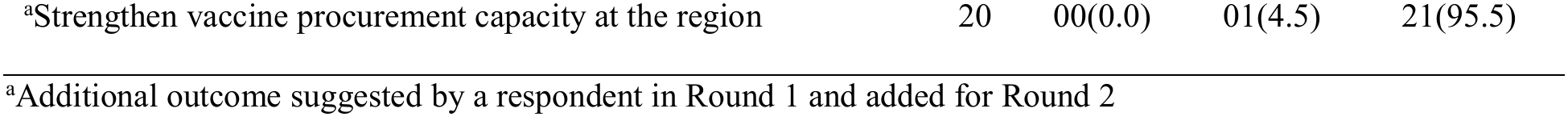
Likert Scale Scores for ‘Strategies to optimize vaccine procurement’ Statements in Round 1.

| **Recommendation** |  | **Round 1** |  |  |
|---|---|---|---|---|
|  |  | **Score** | **Score** | **Score** |
|  |  | **1-3** | **4-6** | **7-9** |
|  | **n** | **n (%)** | **n (%)** | **n (%)** |
| Reinforce community involvement in vaccine procurement | 20 | 02(10.0) | 04(20.0) | 12(60.0) |
| Provision of extra solar refrigerators | 20 | 00(0.0) | 00(0.0) | 20(100.0) |
| Intensified training of staff in data and stock management | 20 | 02(10.0) | 05(25.0) | 13 (65.0) |
| Coordination between districts to reduce vaccine stock-outs | 20 | 02(10.0) | 04(20.0) | 12(60.0) |
| Recruitment of more EPI staff | 20 | 00(0.0) | 02(10.0) | 18(90.0) |
| WHO certification training for EPI staff | 20 | 00(0.0) | 00(0.0) | 20(100.0) |
| Bulk (3 months) requisition for hard to access health districts | 20 | 01(5.0) | 02(10.0) | 17(85.0) |
| Allocation of finance for transport, equipment and training | 20 | 00(0.0) | 00(0.0) | 20(100.0) |
| Regular supervision/monitoring from the region/central level | 20 | 00(0.0) | 02(10.0) | 18(90.0) |
| Provision of transport logistics to district to carry vaccines | 20 | 01(5.0) | 02(10.0) | 17(85.0) |
| In-service training for both district and health facility staff | 20 | 00(0.0) | 00(0.0) | 20(100.0) |
| Support health facilities in vaccine procurement | 20 | 02(10.0) | 04(20.0) | 12(60.0) |
| <sup>a</sup>Resolving the socio-political crisis | 20 | 00(0.0) | 00(0.0) | 20(100.0) |
| <sup>a</sup>Making provision for IDPs during vaccine requisition | 20 | 00(4.5) | 04(20.0) | 16(80.0) |
| <sup>a</sup>Regular evaluation meetings to monitor vaccine coverage | 20 | 00(0.0) | 02(10.0) | 18(90.0) |
| <sup>a</sup>Provision of specialized transport logistics for maritime HDs | 20 | 00(0.0) | 01(4.5) | 21(95.5) |
| <sup>a</sup>Strengthen vaccine procurement capacity at the region | 20 | 00(0.0) | 01(4.5) | 21(95.5) |

<sup>a</sup>Additional outcome suggested by a respondent in Round 1 and added for Round 2

**Table 6:** Framework for the optimization of vaccine procurement.

| Domain | Activity |
|---|---|
| A. Finance | Allocation of finance to health districts for transport, equipment and training |
| B. Workforce | Recruitment of more EPI staff |
|  | WHO certification training for EPI staff |
| C. Transportation logistics | <sup>a</sup>Provision of specialized transport logistics for maritime HDs |
|  | Provision of transport logistics to district to carry vaccines |
| D. Cold Chain Equipment | Provision of (extra) solar refrigerators |
| E. Management/Supervision | Regular supervision/monitoring from the region/central level |
|  | <sup>a</sup>Regular evaluation meetings to monitor vaccine coverage |
| F. In-service training | In-service training for both district and health facility staff |
| G. Vaccine Supply | Bulk (3 months) requisition for hard to access health districts |
|  | <sup>a</sup>Making provision for IDPs during vaccine requisition |
| H. Regional VPC | <sup>a</sup>Strengthen vaccine procurement capacity at the region |
| I. Insecurity | <sup>a</sup>Resolving the socio-political crisis |

<sup>a</sup>Additional outcome suggested by a respondent in Round 1 and added for Round 2

### Priority strategies for the optimization of vaccine procurement

Of the 12 strategies retained for the optimization of vaccine procurement, 5 (41.7%) were priority strategies as seen on figure 2. These strategies include provision of extra solar refrigerators (100.0%), WHO certification training for EPI staff (100.0%), allocation of finance for transport (100.0%), equipment and training, in-service training for both district and health facility staff (100.0%) and resolving the socio-political crisis (100.0%).

**Figure 2:**
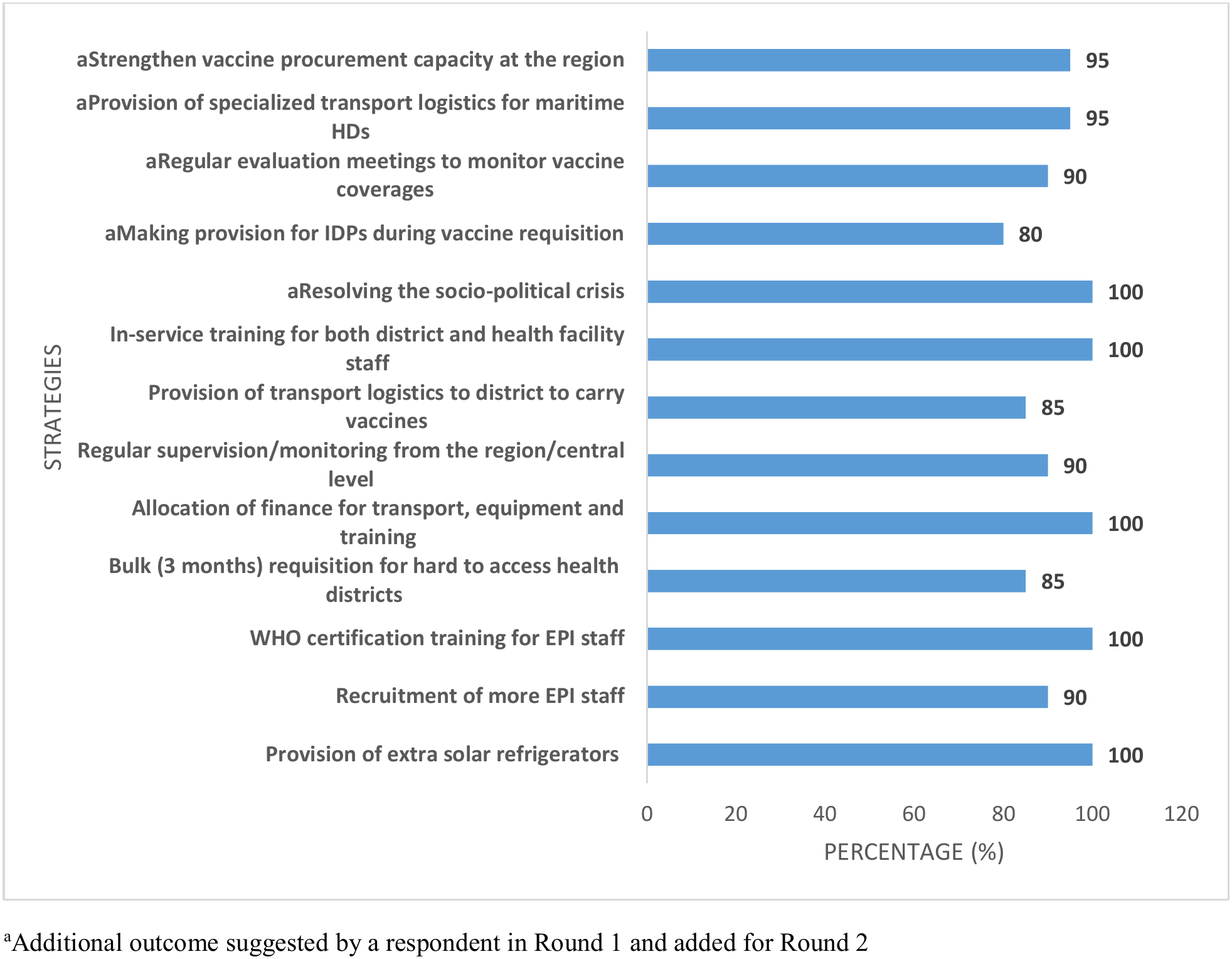
Priority strategies for the optimization of vaccine procurement.

**Figure 3:**
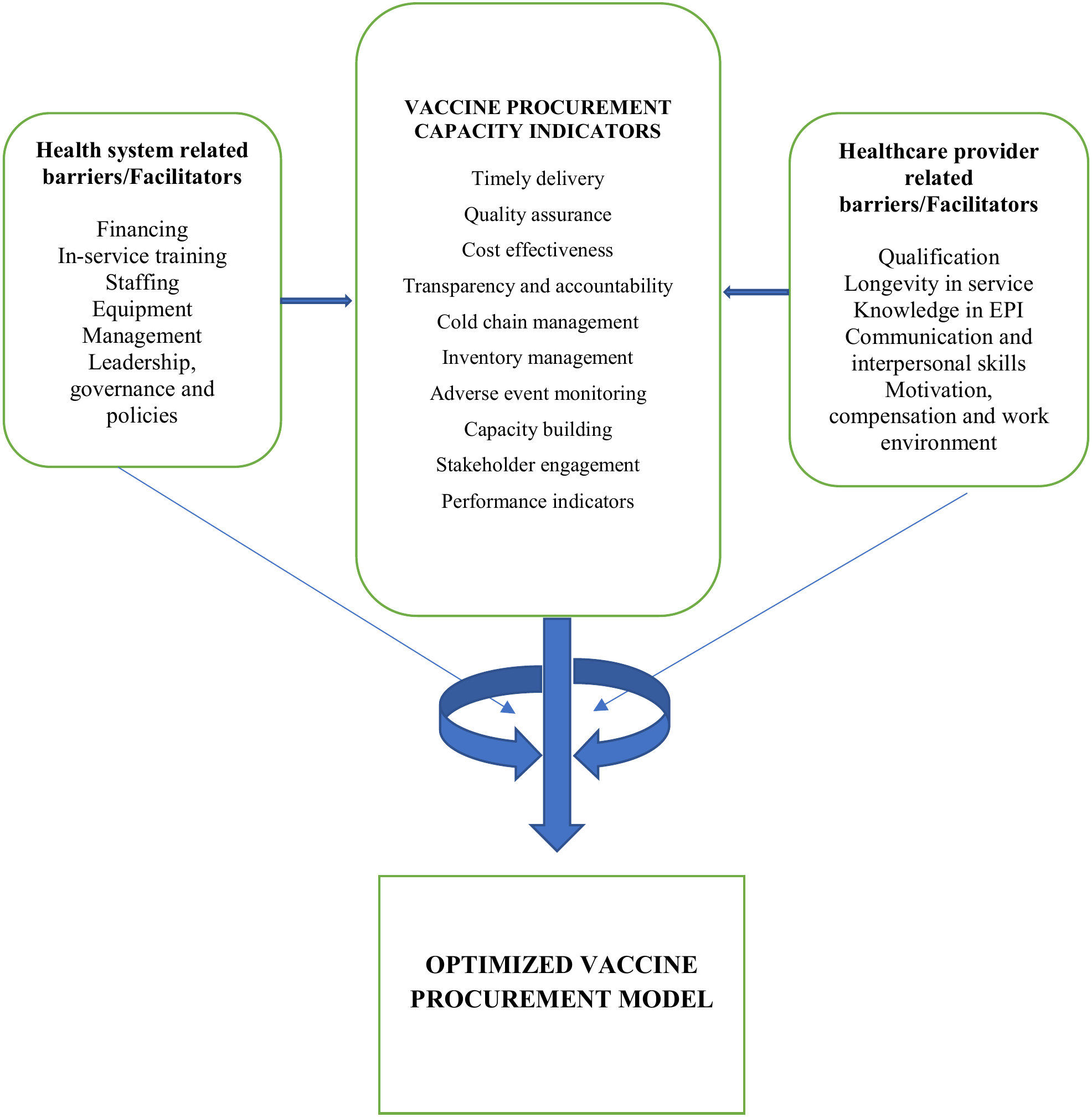
Conceptual Framework for the Optimization of Vaccine Procurement.

### Framework for the optimization of vaccine procurement

The strategies for the optimization of vaccine procurement were grouped into the following domains: finance, workforce, transportation logistics, equipment, management/supervision, in-service training, vaccine supply, regional VPC and insecurity as shown in Table 7.

## DISCUSSION

### Barriers to vaccine procurement in health districts in the South West Region

Our study is one of the few studies that have explored the barriers to vaccine procurement in a conflict zone. The current study is the first to develop strategies for vaccine procurement optimization at the health district level in the conflict-hit zone. Eight broad themes; financing vaccine procurement, recruitment/certification of staff, provision of transportation logistics, provision of solar refrigerators, regular supervision, organize in-service training, training in requisition/stock and bulk-requisition emerged from the thematic analysis of the IDIs. These findings agree with a study in Mozambique, which reported poor coordination and financing as a major bottleneck in vaccine procurement [2]. This highlights the need for streamlining the procurement process and strengthening coordination and collaboration among stakeholders involved. The current study found that majority of the participants said they encountered financial challenges in the course of vaccine procurement since this activity did not have direct funding. They indicated that there are usually no finances allocated to transport the vaccines from the region to the district. The majority of participants from the IDI (all age groups and genders) highlighted that staff shortage was a major hindrance to procurement of vaccines in the south West Region of Cameroon. Some health districts indicated lack of WHO certified cold chain manager and data manager. The absence of these professionals could lead to regular stock-outs and poor vaccine quality assurance.

A few key informants indicated that their cold chains were inadequate as they had faulty refrigerators. This could greatly affect the quality of vaccines sent out to the health districts. In tandem with a study carried out in Cameroon, almost 27.50% of the health facilities were conducting EPI activities without any cold chain equipment [14]. In addition, access to power source was not permanent; cold chain required material such as thermometer and temperature monitoring charts, which were lacking in some health facilities. Activities that are required to ensure cold chain for vaccines such as correct packing of vaccines and ice packs in the cold chain equipment, recording temperature twice a day were not systematically implemented in low-income settings. This increases the risk of exposing vaccine to high temperatures and can seriously hamper the organization of immunization sessions, especially in outreach and mobile strategies. It can also lead to increasing vaccines wastage rates and stock outs. This weakness had been documented in Bongor (Chad) in 2020 [15] and was associated with increase of incidence of measles cases among vaccinated children [15,16]. The situation is expected to be worse in Cameroon if this limited access to cold chain equipment (in 15% of health facilities visited in this study) is combined to intermittent power supply (in 79.4% of health facilities with power supply in the present study).

Irregular power supply of health facilities and absence of standby solar generator were identified in a previous study conducted in Nigeria as one of major barriers to vaccine procurement [17]. It was a positive point to note that for 85.0% of cold chain equipment, temperature-monitoring charts, thermometer, and ice packs were available. This indicates a certain degree of implementation of cold chain monitoring following the national and international EPI Standard Operating Procedures [18]. However, actions are needed to ensure day-to-day monitoring or supervision of the cold chain at the district level. The insufficient supervision of the vaccine procurement process by regional services illustrates failure to implement Standard Operating Procedures for EPI in a considerable proportion of health districts in Cameroon. The absence or insufficient training, supervision, motivation, access to guideline or work load due to lack of health personnel have been highlighted as the major barriers to regular supervision [19]. Similar situation was recently documented in Ethiopia [20, 21] where 29.7% and 12.5% had respectively incomplete and absence of temperature recording on temperature monitoring charts. Some key interventions including constant supervision, training of professionals in charge, promoting access to existing guidelines and making available cold chain tools and equipment in health facilities have been identified to reduce theses gaps [22, 23, 24]. Furthermore, innovating strategies like computerizing temperature monitoring of the vaccine cold chain and developing thermostable vaccines are being tested and could improve the protection of vaccine potency in developing countries context [25, 26, 27].

Some of the key informants indicated that the in-service training on vaccine procurement was inadequate. There were only few training sessions per year (only one session on vaccine procurement in 2025). This could be due to the reduction in foreign aid. Some of the key informants indicated that leading health facilities do not come to collect their vaccines on time at the level of the health district. This could slow down vaccine procurement, cause unnecessary stock-outs, and drop in vaccine coverage. Majority of the key informants reported supervision from the regional level was grossly absent. This could be blamed on the security stakes of the region. The end-point here is that the districts would be left lose to do whatever pleases them. A few key informants indicated that security challenges prevented them from access to the region to collect vaccine and attend in-service training. This had a direct repercussion on vaccine stocks and coverage at the level of the health district. Majority of the key informants reported vaccine stock out especially for polio and HPV due to stock-out at the regional and central levels.

### Framework for the optimization of vaccine procurement in health districts in the South West Region

The current study revealed that of the 12 strategies retained for the optimization of vaccine procurement, 5 (41.7%) were priority strategies (Figure 2). These strategies include: provision of extra solar refrigerators (100.0%), WHO certification training for EPI staff (100.0%), allocation of finance for transport (100.0%), equipment and training, in-service training for district and health facility staff (100.0%), resolving the socio-political crisis (100.0%) and provision of specialized transport logistics for maritime HDs. To solve the vaccine procurement puzzle, study participants made the recommendations shown in Table 3. Some of these recommendations are currently being implemented and they have started to yield positive results, including enabling Cameroon to meet its co-financing requirements for 2030 and purchasing the required quantify of traditional vaccines. The country is also on track to complete its vaccine payment for 2030. Given this earlier success, the authors would like to recommend GAVI and other global organizations to implement a similar approach in other countries. Already, GAVI has pledged to fund the technical working group (TWG) in Cameroon in 2030, and this may be the beginning of helping Cameroon to successfully transition from GAVI support, which will be due in 2027. Given its promise, this approach can help other countries to successfully transition from GAVI support and provide long-term sustainability for the financing of their immunization programs. The concept can also be applied to other vertical programs, including HIV, Malaria and Tb.

## CONCLUSION

The main barriers to vaccine procurement at the level of the health district were financial constraints, limited workforce, inadequate transportation logistics, poor equipment, inadequate managerial skills, inadequate in-service training, vaccine stock-out and weak PHC facilities. The main strategies suggested to overcome these barriers were the financing of vaccine procurement, recruitment/certification of staff, provision of transportation logistics, provision of solar refrigerators, regular supervision, organize in-service training, training in requisition/stock and bulk-requisition.

Based on our findings, we recommend that the Ministry of Public Health should allocate funds for transport, equipment and training on vaccine procurement and also provide solar refrigerators to health districts with interrupted power supply to guarantee a reliable cold chain system.. These will help health districts to limit vaccine stock-out and increase coverage. The Regional Delegation of Public Health should provide WHO certification training for EPI staff and facilitate coordination between health districts in vaccine stock management to reduce vaccine stock-outs. Regular supervision from the central and region level is recommended to give technical support to EPI staff at the district level. Provision of specialized transport logistics for maritime HDs is necessary.

It is imperative for health districts to reinforce community involvement in vaccine procurement. This has a positive bearing on vaccine coverage. More EPI staff should be recruited and trained on data and vaccine stock management to avoid under or over estimation of stock. Anticipation during vaccine requisition is important especially for immunization campaigns and should take IDPs into consideration. Bulk (3 months) requisition should be envisaged for hard to access health facilities / districts and evaluation meetings should be organized regularly to monitor vaccine coverage and requisition. The government of Cameron should strive to resolve the on-going socio-political crisis which the study area. This will facilitate vaccine procurement and coverage.

### Limitations and Strength

The strength of this study is that it involved all the 21 health districts that make up the South West region of Cameroon. In addition, we used the e-Delphi consensus approach (making use of a college of experts in immunization in the South West Region) to identify the best strategies to optimize vaccine procurement capacity of health districts.

## Data Availability

The data that support the findings of this study are available from the corresponding author upon reasonable request.

## ABBREVIATIONS

BCG: Bacillus Calmette -Guerin
CDC: Centre for Disease Control and Prevention
CNS: Central Nervous System
COC: Chief of Center
EPI: Expanded Program on Immunization
GAVI: Global Alliance for Vaccine Initiatives HD- Health District
IDP: Internally Displaced Person
IEC: Information Education and Communication PHC- Primary Health Care
SDG: Sustainable Development Goal
SSA: Sub-Saharan Africa
TWG: Technical Working Group
UNICEF: United Nation International Children’s Fund
VC: Vaccine Coverage
VPD: Vaccine Preventable Diseases
WHO: World Health Organization

## AUTHORS’ CONTRIBUTIONS

FBT: Conceptualization, Data collection, curation, analysis, investigation, methodology, project administration, resources, software, validation, visualization, preparation of manuscript; NFF: conceptualization, investigation, methodology, resources, preparation of manuscript; TAY, ESBT: Data curation, analysis, investigation, methodology, software, preparation of manuscript;, JFA, JFC: conceptualization, methodology, project supervision, validation, and review of manuscript.

## CONSENT FOR PUBLICATION

This manuscript does not contain any individual’s data. Consent for publication was obtained from all the authors who contributed.

## ETHICS APPROVAL AND CONSENT TO PARTICIPATE

The Ministry of Public Health, Regional Ethics Committee for Human Research, South West Region (Ref. No480/CRERSH/SW/C/05/2025), approved this study for implementation.

